# Antiretroviral treatment-induced decrease in immune activation contributes to reduced susceptibility to tuberculosis in HIV-1-TB co-infected persons

**DOI:** 10.1101/2020.11.13.20231274

**Authors:** Katalin A. Wilkinson, Deborah Schneider-Luftman, Rachel Lai, Christopher Barrington, Nishtha Jhilmeet, David M Lowe, Gavin Kelly, Robert J Wilkinson

## Abstract

Antiretroviral treatment (ART) reduces the risk of developing active tuberculosis (TB) in HIV-1 co-infected persons. In order to understand host immune responses during ART in the context of Mycobacterium tuberculosis (Mtb) sensitization, we performed RNAseq analysis of whole blood-derived RNA from HIV-1 infected patients during the first 6 months of ART. A significant fall in RNA sequence abundance of the Hallmark IFN-alpha, IFN-gamma, IL- 6/JAK/STAT3 signaling, and inflammatory response pathway genes indicated reduced immune activation and inflammation at 6 months of ART compared to day 0. Further exploratory evaluation of 65 soluble analytes in plasma confirmed the significant decrease of inflammatory markers after 6 months of ART. Next, we evaluated 30 soluble analytes in QuantiFERON Gold in-tube (QFT) samples from the Ag stimulated and Nil tubes, during the first 6 months of ART in 30 patients. There was a significant decrease in IL-1alpha and IL-1beta (Ag-Nil) concentrations as well as MCP-1 (Nil), supporting decreased immune activation and inflammation. At the same time, IP-10 (Ag-nil) concentrations significantly increased, together with chemokine receptor-expressing CD4 T cell numbers. Our data indicate that ART-induced decrease in immune activation combined with improved antigen responsiveness may contribute to reduced susceptibility to tuberculosis in HIV-1-TB co-infected persons.

## Introduction

Tuberculosis (TB) remains the leading bacterial cause of death worldwide (1). The biggest recognised risk factor for developing TB disease is Human immunodeficiency virus (HIV-1) infection (2). Antiretroviral treatment (ART) is an effective way to reduce the risk of TB in HIV- 1 co-infected persons, reducing TB incidence between 54-92 % at the individual level (3). More broadly, ART was shown to be associated with the decline of TB in sub-Saharan Africa between 2003-2016, preventing an estimated 1.88 million cases (4). In order to understand how ART reduces susceptibility to TB in HIV-1 infected persons, we longitudinally analysed a group of HIV-1 infected persons over 6 months from starting ART (day 0). Our hypothesis was that this highly susceptible group who undergo immune reconstitution through ART, and thereby become less susceptible to TB, will yield insight into protective mechanisms against TB in humans.

Before the commencement of ART, the risk of TB is increased at all stages of HIV-1 infection. This could be due to depletion of *Mycobacterium tuberculosis* (Mtb) specific T cells early during HIV-1 infection (5) as well as impairment of function of these antigen-specific CD4 T cells (6). We and others have shown that increased ART-mediated immunity broadly correlates with the expansion of early differentiated (central memory) T-cell responses and overall reduction in cellular activation (7-9). More specifically, we recently demonstrated that the numbers of Mtb-antigen specific CD4 T cells increase during the first 6 months of ART, together with proportionally expanded polyfunctionality (cells co-producing TNF, IFN-gamma and IL-2), and a concomitant decrease in their activation profile (10).

As the risk and incidence of TB remains much higher in HIV-1 co-infected persons compared to those not infected with HIV-1, despite widespread implementation of ART regimes globally and even during long-term ART (11), it is important to understand the host immune response in the context of Mtb infection and ART. Therefore, we performed exploratory RNAseq analysis of whole blood derived RNA from HIV-1 infected patients during the first 6 months of ART, followed by evaluation of multiple soluble analytes in plasma. For more precise assessment of the change in soluble plasma biomarkers in the context of Mtb sensitisation during ART, we used QuantiFERON® Gold in-tube (QFT) plasma samples. These samples have been shown to be useful in evaluating host biomarkers other than IFN-gamma, regardless of HIV-1 status (12) (13). Moreover, we recently used similar QFT plasma samples to evaluate the predictive performance of 13 plasma biomarkers in HIV-1 infected individuals likely to progress to active TB and found that unstimulated plasma analyte concentrations better identified TB risk in these HIV-1 co-infected patients, demonstrating that underlying inflammatory processes and higher overall background immune activation might render them more susceptible to progress to TB (14). However, no studies so far evaluated QFT plasma analyte concentrations longitudinally, during ART. Here we present data suggesting that the ART- induced decrease in immune activation combined with improved antigen responsiveness may contribute to reduced susceptibility to tuberculosis in HIV-TB co-infected persons.

## Results

### 1. RNA sequencing analysis

Exploratory analysis performed in six samples from D0 and six samples from 6M (not paired) revealed 155 differentially expressed genes (Figure 1A, Supplementary Figure 1A and Supplementary Table 2). Gene Set Enrichment Analysis (GSEA) using Reactome identified a significant fall in the Interferon-alpha, Interferon-gamma, IL-6/JAK/STAT3 signaling and overall inflammatory response pathway genes (Figure 1B). These results were replicated by similar findings using Hallmark pathway analysis as well (Supplementary Figure 1B). While interestingly there was an increase in Heme metabolism as shown by GSEA (Figure 1B), overall the data points towards reduced immune activation and inflammation at 6 months of ART compared to day 0.

**Figure 1.**
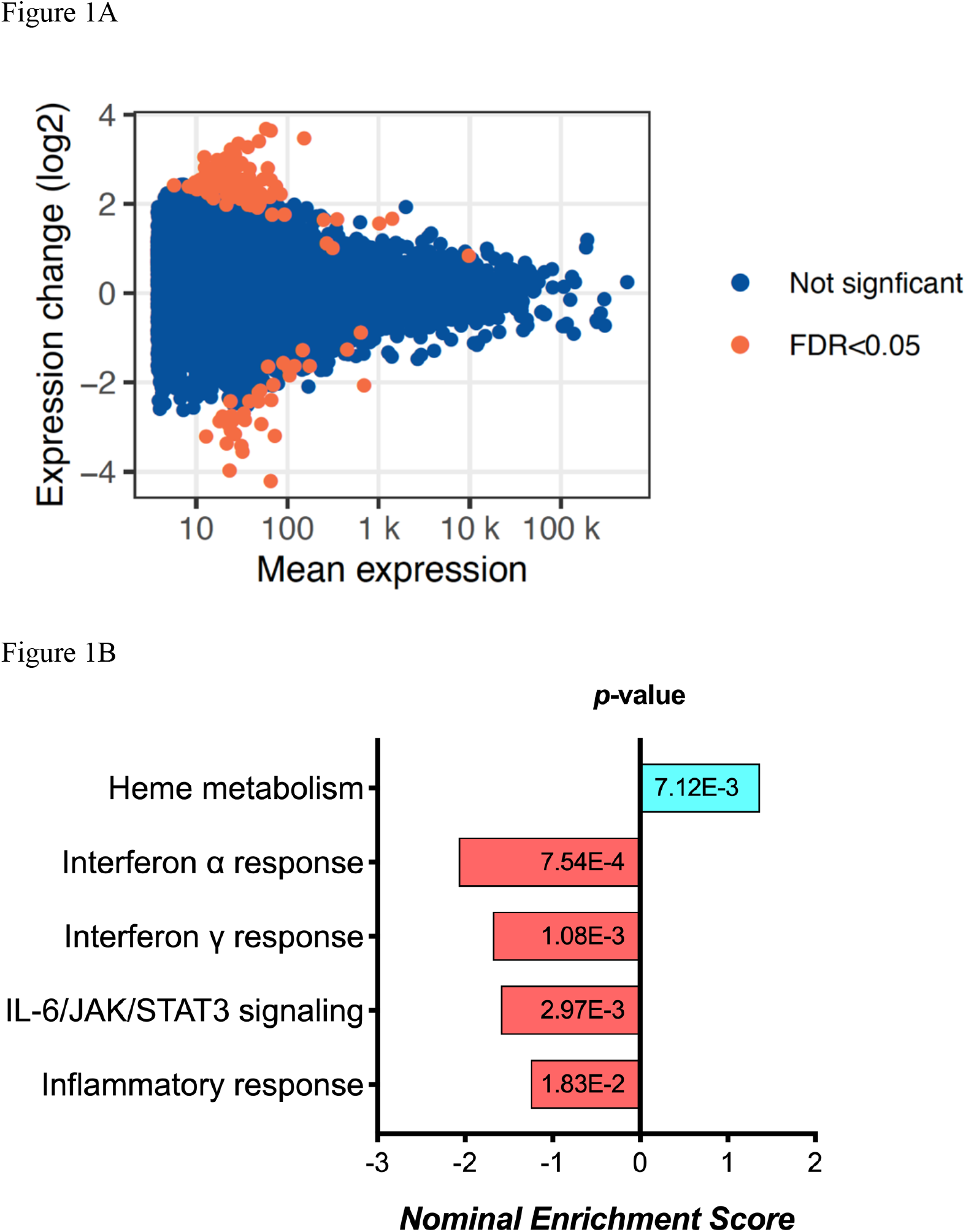
RNA sequencing analysis results, showing (A) the MA plot visualising the statistically different genes by transforming the data onto M (log ratio) and A (mean average) scales; and (B) the pathways significantly changed between day 0 and 6 months of ART based on Gene set enrichment analysis (GSEA).

### 2. Multiplex analysis using plasma

Next we used plasma collected at day 0 and 6 months of ART (n=39, paired samples) in further exploratory analysis of soluble markers using the Immune Monitoring 65-Plex Human ProcartaPlex™ Panel (Invitrogen, Thermo Fischer). 29 analytes were excluded from analysis due to (1) being undetectable in all samples (FGF-2, IL-5, IL-31, MCSF, MIF, TNF-beta), 99% of samples (MCP-3, IL-23), 96% of samples (IL-7); (2) the median concentration being 0 pg/ml at both timepoints (bNGF, G-CSF, GM-CSF, GRO-alpha, IL-1alpha, IL-1beta, IL-3, IL-4, IL-6, IL- 8, IL-9, IL-10, IL-12p70, IL-15, IL-20, IL-22, IL-27, LIF); (3) the median concentration being 1 pg/ml at both timepoints, although there was a trend showing decrease over time (IFN-alpha, IL- 13). The remaining results are summarized in Table 1, demonstrating a significant decrease in soluble plasma analytes overall. Since these plasma analyte determinations were exploratory, we did not perform correction for multiple comparisons. However, we did utilize an alternative immune marker monitoring platform, Meso Scale Discovery system, to further confirm the trends seen in selected markers from all groups (highlighted by italics in Table 1). Results are summarized in Supplementary Table 3. While there were notable differences in the range of concentrations detected by the two different platforms, as shown previously by others (15-18), overall the data confirm the significant decrease in HIV-1 induced immune activation and inflammation over the first 6 months of ART. Since the MSD Angiogenesis panel allows individual evaluation of VEGF-A, C, and D, it was interesting to find an increase over time in VEGF-D plasma concentrations (from median of 851 to 1046 pg/ml, p=0.0002, Supplementary Table 3).

**Table 1.** Multiplex analysis of plasma.

| nr | Analyte<sup>1</sup> | day 0 median | day 0 (IQR) | 6M median | 6M (IQR) | p value<sup>2</sup> |
| --- | --- | --- | --- | --- | --- | --- |
|  | **T cell response related** |  |  |  |  |  |
| 1 | *IFN-gamma* | 53 | 34.1-80.3 | 27 | 19.8-37.7 | <0.0001 |
| 2 | IL-18 | 116 | 30.5-187.6 | 51 | 18.5-104.3 | <0.0001 |
| 3 | CD40-Ligand | 236 | 0-305 | 0 | 0-245 | <0.0001 |
| 4 | *IP-10* | 240 | 109-375 | 67 | 24-148 | <0.0001 |
| 5 | MIG | 968 | 707-1256 | 526 | 0-715.5 | <0.0001 |
| 6 | IL-16 | 1637 | 1107-1840 | 1119 | 743-1610 | <0.0001 |
| 7 | IL-2R | 25084 | 15178-40542 | 7953 | 4746-13525 | <0.0001 |
| 8 | TSLP | 19 | 9.9-30.5 | 4 | 0-15 | 0.0006 |
| 9 | I-TAC | 314 | 0-735 | 0 | 0-326 | 0.0014 |
| 10 | IL-2 | 32 | 11.5-91.1 | 18 | 11.5-106.7 | 0.19 |
| 11 | IL-21 | 98 | 0-222.3 | 62 | 0-200.4 | 0.1 |
| 12 | *IL-17A* | 348 | 56-690 | 309 | 135.7-674.5 | 0.26 |
|  | **TNF superfamily related** |  |  |  |  |  |
| 13 | TNF-RII | 662 | 585-717 | 569 | 521-633 | <0.0001 |
| 14 | Tweak/TNFSF12 | 2266 | 1286-3432 | 1306 | 909-1779 | <0.0001 |
| 15 | CD30 | 10436 | 7935-14690 | 4673 | 3001-6851 | <0.0001 |
| 16 | ENA-78(LIX) | 859 | 407-1973 | 603 | 323-1425 | 0.004 |
| 17 | APRIL | 4274 | 3390-5268 | 3836 | 2893-4846 | 0.005 |
| 18 | *TNF-alpha* | 3 | 1.48-7.6 | 3 | 0.35-4.06 | 0.027 |
| 19 | TRAIL | 682 | 0-966 | 0 | 0-804 | 0.034 |
| 20 | BAFF | 49 | 0-96.7 | 0 | 0-126.6 | 0.427 |
|  | **Monocyte / macrophage related** |  |  |  |  |  |
| 21 | MCP-2 | 25 | 12.9-38.9 | 11 | 6.6-15.7 | <0.0001 |
| 22 | *MIP-1alpha* | 17 | 13-24 | 10 | 6.1-15 | <0.0001 |
| 23 | BLC | 1113 | 593.5-2170 | 422 | 155.7-755.8 | <0.0001 |
| 24 | Eotaxin-3 | 72 | 0-102.8 | 0 | 0-36.7 | 0.0002 |
| 25 | MDC/CCL22 | 4374 | 1529-10229 | 1515 | 446-4264 | 0.0004 |
| 26 | MIP-3alpha | 26 | 9.7-55.9 | 10 | 2.7-35.1 | 0.0016 |
| 27 | *MIP-1beta* | 61 | 30.2-92.2 | 42 | 14.1-63.4 | 0.008 |
| 28 | Eotaxin-2 | 6 | 0.9-20.3 | 2 | 0-12.3 | 0.071 |
| 29 | Fractalkine | 16 | 9.5-29.9 | 12 | 5.5-23 | 0.074 |
| 30 | *MCP-1* | 253 | 121.7-388.2 | 211 | 104.6-319.7 | 0.22 |
| 31 | Eotaxin-1 | 14 | 7.3-38.7 | 11 | 5.6-47.9 | 0.78 |
|  | **Growth factors** |  |  |  |  |  |
| 32 | *VEGF-A* | 815 | 512.7-1055 | 425 | 223.2-658.8 | <0.0001 |
| 33 | HGF | 565 | 427.7-714 | 456 | 329-502.6 | 0.0005 |
| 34 | MMP-1 | 1879 | 1065-4429 | 1649 | 764-2731 | 0.049 |
| 35 | SDF-1alpha | 7690 | 6142-8754 | 6940 | 5496-8884 | 0.085 |
| 36 | SCF | 38 | 23.1-48.8 | 34 | 17.4-48.8 | 0.59 |

<sup>1</sup> analyte concentrations expressed as median (IQR) pg/ml; <sup>2</sup>Wilcoxon matched pairs (n=39).

### 3. Multiplex analysis using QFT plasma

In order to assess more precisely the change in soluble plasma biomarkers in the context of Mtb sensitisation during ART, we next used QuantiFERON® Gold in-tube (QFT) plasma samples. Here we longitudinally analyzed 30 soluble analytes in 30 HIV-1-infected individuals (9 male, 21 female; mean age 34.3 years) at baseline (D0) and after one (1M), three (3M) and six months (6M) of receiving ART, using the Nil and selected Antigen-stimulated (Ag-Nil) QFT plasma samples. The HIV-1 viral load (VL) and CD4 counts of the 30 participants during ART are summarized in Supplementary Table 4.

First, we assessed the effect of antigen stimulation at each timepoint by comparing Ag stimulated samples to unstimulated (Nil) samples. The QFT assay was optimized to assess an IFN-gamma response in the form of (Ag-Nil), however, here we intended to assess the effect of antigen stimulation on other analytes as well and in order to be able to compare the effects we opted to assess the ratio, or fold change in analyte concentration between Ag stimulated and Nil assays. The analytes that showed a significant response difference over time are summarized in Table 2 and were selected to be included in further analysis in the form of (Ag-Nil). All data related to the analysis are summarized in Supplementary Table 5.

**Table 2.**
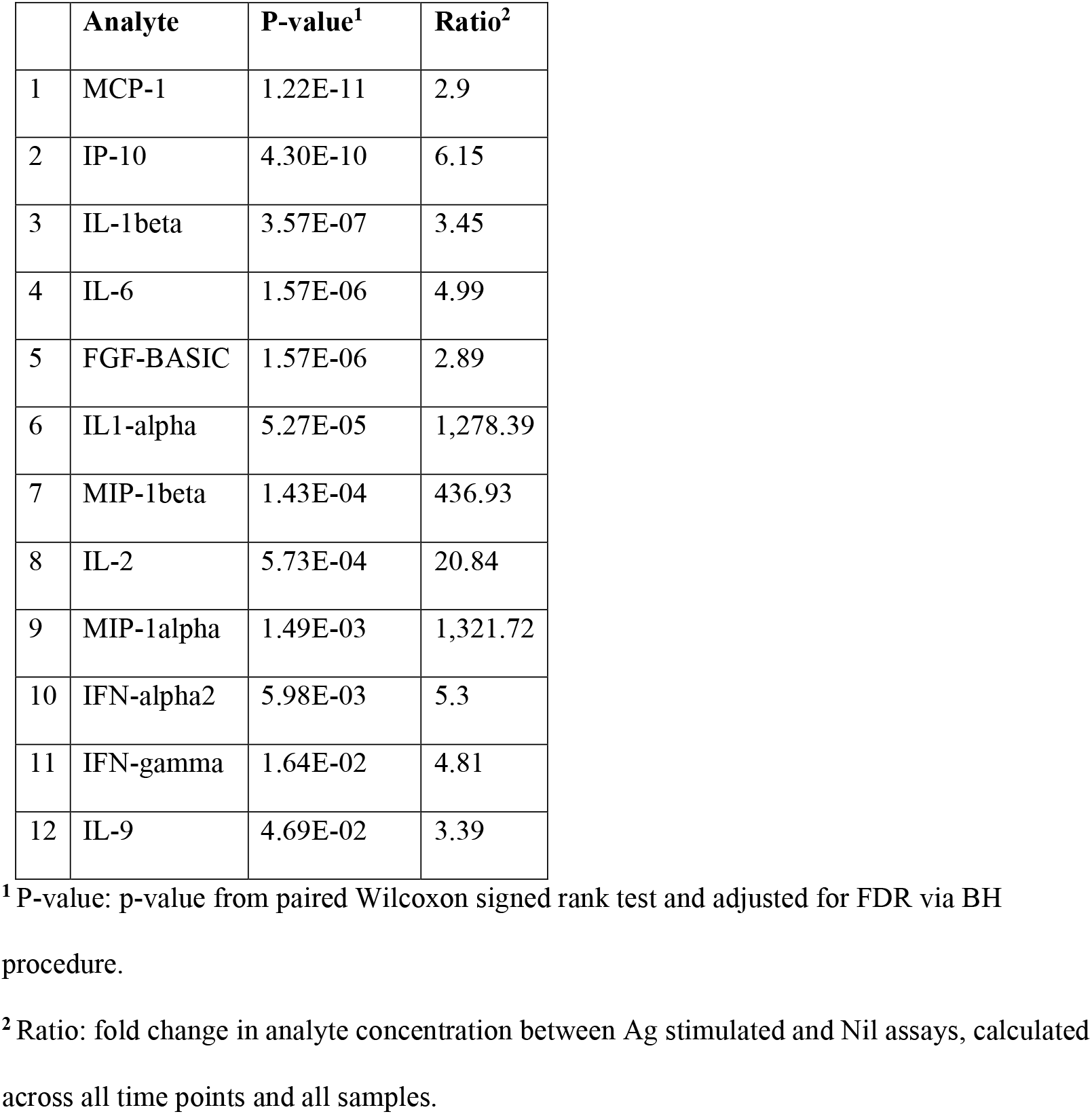
Analytes found to be significantly changed by Ag stimulation, that were retained for further analysis.

The change in analytes at 1 month, 3 months and 6 months of ART, compared to day 0 is shown in Supplementary Figure 2, while the consistent change in analytes across all time points is summarized in Figure 2. There was a consistent decrease in IL-1alpha and IL-1beta concentrations in the (Ag-Nil) samples, as well as MCP-1 concentration in the Nil samples (as already seen in the plasma analyte evaluation above using MSD). Interestingly, there was an increase in antigen specific (Ag-Nil) IP-10 concentrations, especially at 6 months of ART. The fold change in these analyte concentrations at each timepoint is shown in Table 3. IL-1beta and IL-1alpha (Ag-Nil) were also found to be strongly correlated (corr = 0.55, p= 4.14e-06) at 6 months, but not correlated to IP10 (corr = −0.075, p=0.56 for IL-1alpha, corr = - 0.130, p=0.33 for IL-1beta). This is also reflected in the principal components analysis (PCA) loading plot (Figure 3A showing the first two principal components), where IL-1alpha and IL- 1beta strongly influence PC1 (53% of variance), while IP-10 almost solely, drives PC2 (32% of variance). This was supported by clustering analysis using K-means, that found K=3 clusters to be optimal for the data (Figure 3B); with IL-6 (Ag-Nil), MIP-1alpha (Ag-Nil) and IL-8 (Nil) grouped together, IP-10 (Ag-Nil) in its own cluster, and all other compounds grouped in the third cluster (PC1, first principal component, variance explained: 77.4% and PC2, second principal component, variance explained: 18.6%).

**Table 3.** Analytes with significant changes from baseline, over the follow up period.

| analyte<br>(condition) | **Month 1<sup>1</sup>** | **Month 3<sup>1</sup>** | **Month 6<sup>1</sup>** | **Adj. P-Val<sup>2</sup>** |
| --- | --- | --- | --- | --- |
| IL-1beta<br>(Ag-Nil) | -3,240.28 | -4,086.82 | -3,047.68 | 1.43E-03 |
| IL-1alpha<br>(Ag-Nil) | -2,053.01 | -2,791 | -2,283.65 | 2.68E-03 |
| MCP-1<br>(Nil) | -1,152.4 | -1,263.36 | -791.29 | 2.68E-03 |
| IP-10<br>(Ag-Nil) | 22,221.56 | 929.38 | 47,674.16 | 1.61E-02 |

<sup>1</sup> log-fold change between the concentration at baseline (day 0) and concentrations at 1, 3, and 6 months of ART respectively (negative value means decrease and positive value mean increase)
<sup>2</sup> Adj. P-Val: P value from Limma analysis, after adjustment for FDR.

**Figure 2.**
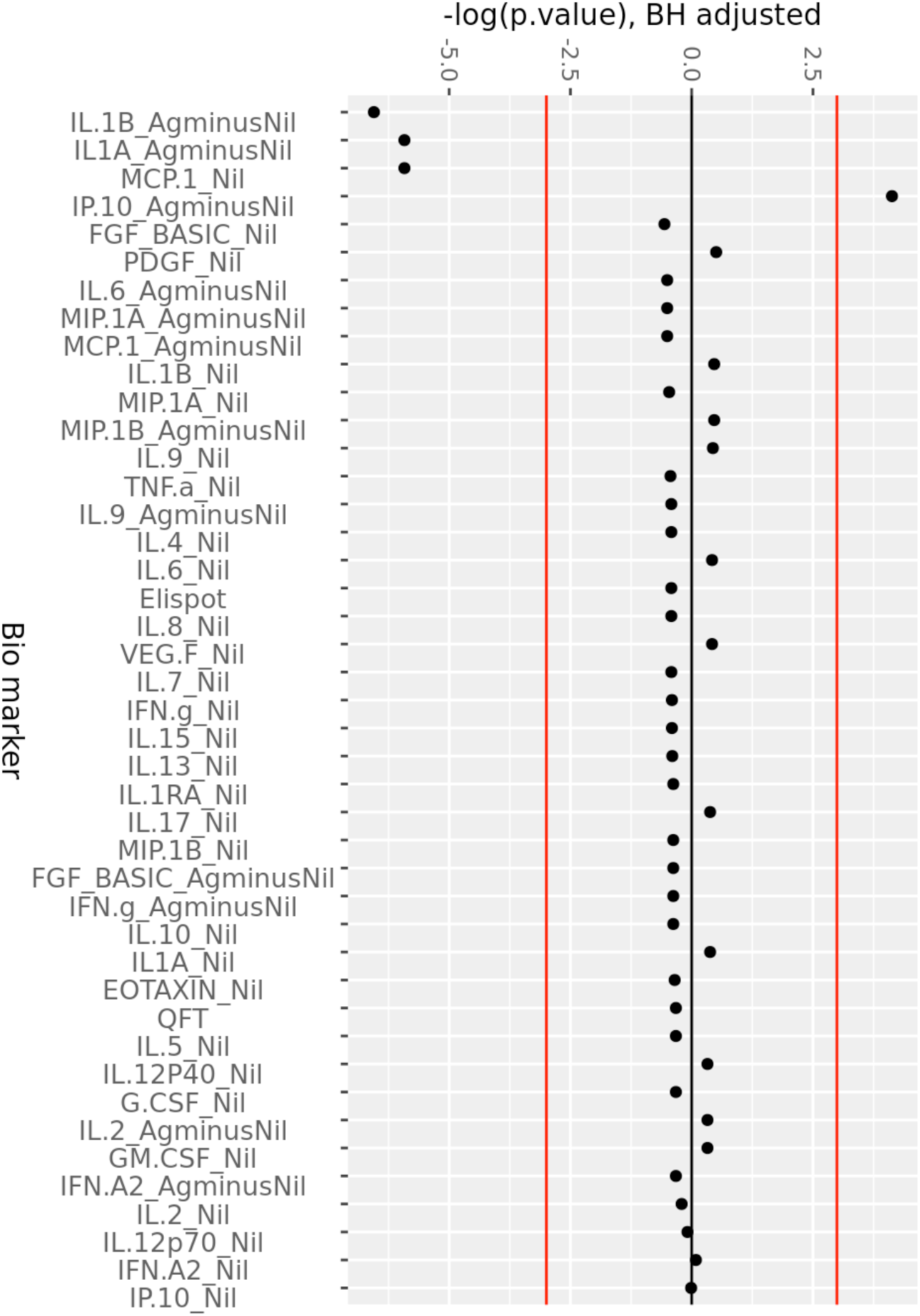
Consistent change in QFT plasma analytes over 6 months of ART. Minus log-transformed p- values for each analyte, resulting from the Limma analysis framework, after BH FDR correction, considering all timepoints. Sign represents direction of effect size (negative: decrease from D0 level, positive: increase from D0 level). Red lines: α significance thresholds.

**Figure 3.**
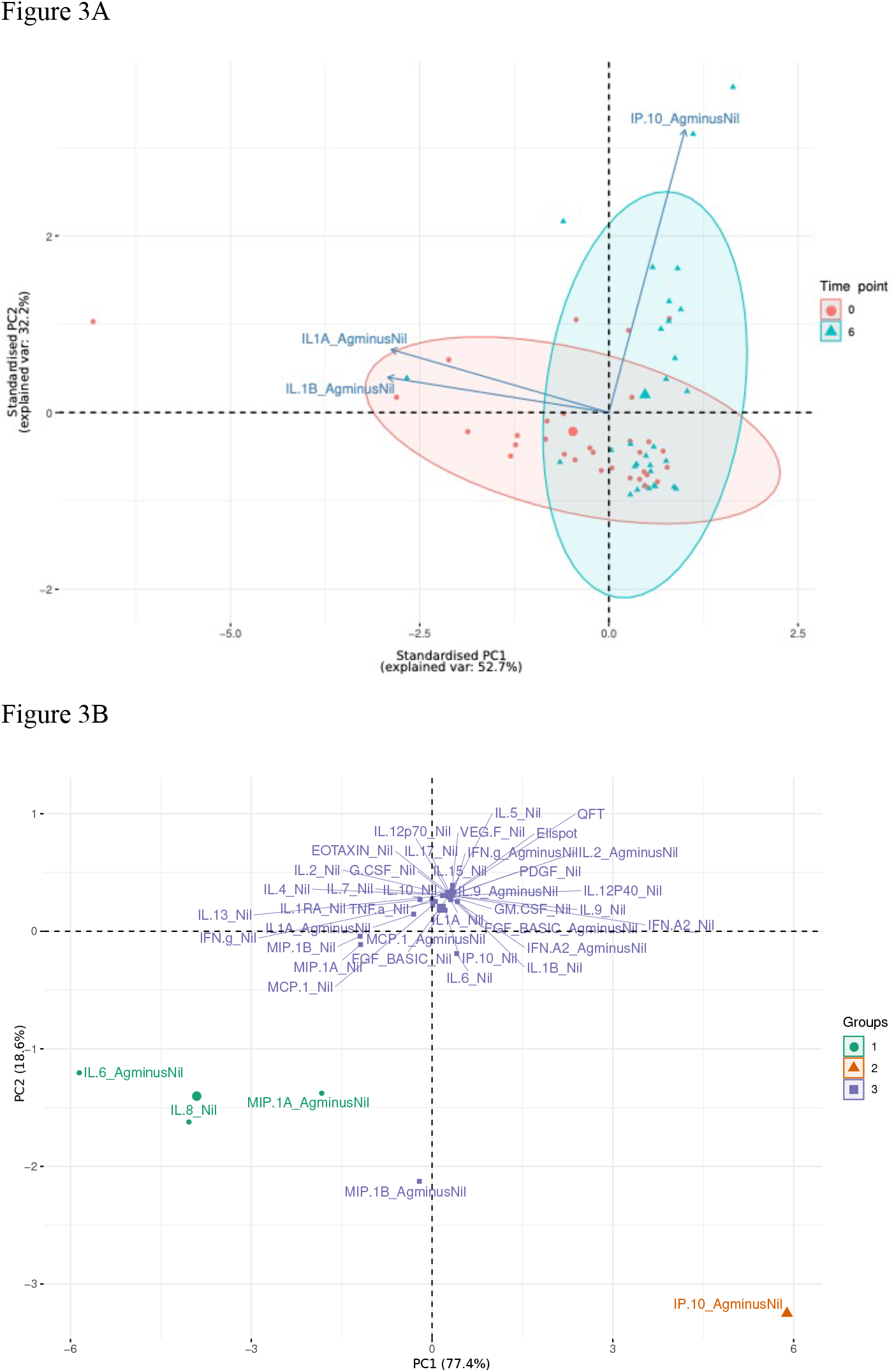
First two principal components plots: (A) Over three most significantly changing analytes (arrows) between day 0 (red) and 6 months (blue), each dot representing a patient sample. (B) Over each analyte of interest and all time points, grouped by K-means clustering, using K=3, each dot representing a compound.

### 4. Chemokine receptor analysis using flow cytometry

Overall, the above data support a decrease in HIV-1 induced immune activation and inflammation over the first 6 months of ART, even in the context of Mtb sensitisation. However, to better understand the increase in antigen specific IP-10 (Ag-Nil) concentrations, we evaluated the expression of chemokine receptors CXCR3, CCR4 and CCR6 on the surface of CD4 T cells, using PBMC in a subset of 25 patients from the same cohort, as previously described (10). We found expanding numbers of CD4 T cells expressing CXCR3 (the receptor for IP-10), as well as CCR4 and CCR6 between day 0 and 6 months or ART. Thus, the number of CD4^+^CXCR3^+^ T cells increased from median 20 (IQR 12-41) cells/μL at day 0 to 23 (IQR 12-68) at 6 months of ART (p=0.009). Similarly, the number of CD4^+^CCR4^+^ T cells increased from median 31 (IQR 6- 40) to 52 (IQR 33-58, p=0.0001) and the number of CD4^+^CCR6^+^ T cells increased from median 27 (IQR 18-35) to 36 (IQR 22-44, p=0.045) respectively, all Wilcoxon matched pairs comparisons for n =25 at day 0 and 6 months of ART, with data shown in Supplementary Figure 3. These results indicate expanding numbers of CD4 T cells that are capable to respond to chemokine signaling over the first 6 months of ART.

## Discussion

The aim of this study was to better understand host immune responses during ART. Our exploratory RNAseq analysis indicates a significant fall in the Hallmark Interferon alpha, Interferon gamma and IL-6-JAK-STAT-signalling, together with overall inflammatory response pathway genes between day 0 and 6 months of ART, suggesting reduced immune activation and inflammation at 6 months of ART compared to day 0. This trend is supported by decreasing concentrations of soluble markers in plasma from the same cohort, indicating an overall decline in HIV-1 induced immune activation and inflammation during the first 6 months of ART. Our more targeted analysis of soluble plasma biomarkers in the context of Mtb sensitisation during ART, using QFT supernatants, shows a decrease in antigen specific IL-beta and IL-1alpha as well as MCP-1, in line with decreased HIV-1 induced immune activation and inflammation. At the same time, antigen specific IP-10 (Ag-nil) concentrations significantly increased, together with chemokine expressing CD4 T cells, in keeping with memory T cell expansion as we demonstrated previously (7, 10). Overall, our data indicates that the ART- induced decrease in immune activation combined with improved antigen responsiveness may contribute to reduced susceptibility to tuberculosis in HIV-1-TB co-infected persons.

The increase in antigen specific IP-10 (Ag-Nil) coincides with increasing numbers of CXCR3+CD4+ T cells. CXCR3, the receptor for IP-10 (also known as IFN-gamma inducible protein 10 or CXCL10), is a chemokine receptor highly expressed on effector T cells and plays an important role in T cell trafficking, effector T cell recruitment from the lymphoid tissue to the peripheral sites and an important role in memory CD4 T cell function throughout the activation and differentiation pathway (19). Our data showing a decrease in plasma IP-10 but an increase in antigen specific IP-10, together with expanding numbers of CD4 T cells expressing various chemokine receptors, are in line with the overall memory CD4 T cell expansion (10) and point towards broadly improved capacity to respond to new infections through better responsiveness to antigen, as a result of ART.

The increase in Heme metabolism as indicated by RNAseq, together with increasing concentrations of VEGF-D in plasma, may reflect reversal of the endothelial dysfunction associated with both HIV-1 infection and ART (20). The heme oxygenase system acts as a potent antioxidant, protects endothelial cells from apoptosis, is involved in regulating vascular tone, attenuates inflammatory response in the vessel wall, and participates in angiogenesis and vasculogenesis (21). Vascular endothelial growth factor-D (VEGF-D) is a secreted protein that can promote the remodeling of blood vessels and lymphatics in development and disease (22). While their increase could indicate normalisation of endothelial integrity and activity on ART, recent evidence suggests that HIV-1 associated endothelial activation persists despite ART (23). Due to increased rates of cardiovascular disease among people living with HIV-1, this is an important area to explore in further studies.

Multiple studies have used QFT plasma samples mainly to differentiate between active and latent TB (24). Our previous analysis of similar samples to identify TB risk in HIV-1 co-infected patients indicated that unstimulated plasma (Nil) analyte concentrations may reflect underlying inflammatory processes and this higher overall background activation might render these co-infected individuals more susceptible to progress to TB (14). We also showed that the activation profile of Mtb-specific CD4 T cells in terms of HLA-DR expression identifies TB disease activity irrespective of HIV-1 status (25). Recent data using the macaque model of SIV/Mtb co-infection indicates that SIV induces chronic immune activation, leading to dysregulated T cell homeostasis which is associated with reactivation of LTBI (26, 27). This may imply that HIV-1 induced chronic immune activation leads to LTBI reactivation and therefore increased susceptibility to TB in HIV-1 co-infected individuals. Taken together, our data supports that ART reduces chronic immune activation, thereby leading to reduced susceptibility to TB, through an overall decrease in HIV-1 induced immune activation and inflammation, including decreased T cell activation, increased numbers of polyfunctional antigen specific T cells (10), with improved capacity to respond to new infections through better responsiveness to antigen.

## Methods

### Study Cohort

HIV-1 infected persons starting ART were recruited from the Ubuntu Clinic in Khayelitsha, South Africa, during two longitudinal studies in 2011-2012. Blood for plasma separation, Quantiferon Gold In-tube (QFT) assay and RNA extraction (Tempus™ tubes) was collected at baseline (Day 0) and after one (1M), three (3M) and six months (6M) of receiving ART. All samples were stored at −80°C for future use. The University of Cape Town Faculty of Health Sciences Human Research Ethics Committee approved these studies (HREC 245/2009 and 545/2010). All participants gave written informed consent in accordance with the Declaration of Helsinki. The two cohorts have previously been described (28-30). All patients experienced an increase in CD4 counts and a decrease in HIV viral load during the first 6 months of ART and were all sensitised by Mtb (as determined by positivity in at least in one interferon gamma release assay, QFT and/or ELISpot, at least one timepoint during the longitudinal follow up) as previously shown (30).

### RNA sequencing library preparation

Total RNA was extracted from whole blood using the Tempus Spin RNA Isolation kit (Thermofisher) according to the manufacturer’s recommendations as described (30). The quantity and quality of the extracted RNA were measured by the Qubit fluorometer and the Caliper LabChip system, respectively. RNA libraries for whole blood RNA were constructed using the Ovation Human Blood RNAseq Library Systems (Tecan, Mannedorf, Switzerland) where ribosomal and globin RNA were removed according to the manufacturer’s protocol. The final libraries were assessed using TapeStation 2200 System (Agilent, Santa Clara, CA). All libraries were sequenced on Illumina Hi-Seq 4000 instrument with paired-end 100 cycle reactions and 40 million reads per sample.

### RNAseq data analysis

Indexed libraries were pooled and sequenced on an Illumina HiSeq 4000 configured to generate 101 cycles of paired-end data. Raw data was demultiplexed and FastQ files created using bcl2fastq (2.20.0). Datasets were analysed using the BABS-RNASeq Nextflow (31) pipeline developed at the Francis Crick Institute. The GRCh38 human reference genome was used with the Ensembl release-86 (32) gene annotations. Dataset quality and replication was validated using FastQC (0.11.7, Andrews, <u>link</u>), RSeQC (33), RNAseqC (34) and Picard (2.10.1, <u>link</u>). Reads were then aligned to the genome and expression quantified using STAR (35) and RSEM (36). Estimates of gene expression were loaded into R version 3.5.1 using the tximport package. DEseq2 (37) was used to test for significant differences between day 0 and 6 months of ART. Default parameters were applied and false discovery rate was set to 0.1. Gene set enrichment analysis was performed using fgsea (38). Hallmark pathways (‘h.all.v6.2.symbols.gmt’) were downloaded (http://software.broadinstitute.org/gsea) and used to test for enrichment genes ranked by their stat value calculated by DESeq2. The data was deposited to Gene Expression Omnibus (GEO) under accession number GSE158208.

### Multiplex immune assays for cytokines and chemokines

The preconfigured multiplex Human Immune Monitoring 65-plex ProcartaPlex immunoassay kit (Thermo Fisher Scientific, UK) was used to measure 65 protein targets in plasma. Bio-Plex Pro Human Cytokine 27-Plex Immunoassay (Bio-Rad Laboratories, Hercules, CA, USA) and customized MilliplexTM kits (Millipore, St Charles, MO, USA) were used to measure 30 analytes in the QFT plasma samples from unstimulated (Nil) and TB specific antigen stimulated (Ag) samples. All assays were conducted on the Bio-Plex platform (Bio-Rad Laboratories, Hercules, CA, USA), using Luminex xMAP technology. Additional confirmatory experiments were run on the Meso Scale Discovery Inc (Rockville, MD) platform, using the plasma samples only (not QFT). All assays were conducted as per the manufacturer’s recommendation and all analytes measured are listed in Supplementary Table 1.

### Flow cytometry analysis

Cryopreserved peripheral blood mononuclear cells were thawed, counted, rested and stained with LIVE/DEAD® Fixable Near-IR Stain and the following surface markers: CD14 and CD19 (APC-AF750), CD4 (FITC), CD45RA (BV570), CD27 (BV711), CXCR3 (PE-Cy7), CCR4 (BV510), CCR6 (BV605). Cells were acquired on a BD-Fortessa flow cytometer and analysed using FlowJo as previously described (10).

### Data cleaning and analysis for QFT plasma samples

After removal of missing data 30 analytes measured at four time-points over the course of 6 months were evaluated: baseline (D0), after one (1M), three (3M) and six months (6M) of receiving ART. Subjects with more than 50% of data missing across all the time points for the analytes of interest were removed. Missing data points (0.8% of all data) were imputed using KNN imputation (k=15) if they were not dropped from the data set. All analytes were sampled in Nil and Ag (antigen stimulated) assays. Ag stimulated assays found to be significantly different than their Nil counterparts were retained for the main analysis, after transformation into (Ag-Nil). These selected (Ag-Ni)] assays and Nil-only assays were considered for the remaining part of the analysis.

### Statistical Analyses for QFT plasma samples

All analyses were carried out in R version 3.6.1, using the analytical framework provided in the Limma package (39). Limma follows a parametric empirical Bayes model within multiple linear regressions. A linear regression model is fitted for each molecule of interest, the resulting estimates are then linked to global hyper-parameters in order to share information between the molecules. This allows to calculate correlations between analytes of interest, together with correlations between related samples, but also helps correct for unequal quality / unequal variance between time points. Limma is also equipped with mixed effect modeling with unconditional growth and constant intra-block correlation, which allows us to adjust for the potential batch effect of data collection at each time point.

Using Limma with mixed effect for each biomarker n ∈[1,N],N= 43, we have:

- *Yn,i,j =β*0,*n,j*+*β*1,*n,jti,j*+*ϵn,i,j*,
- *β*0,*n*,j =*γ*0,*n*+*U*0,*n,j*
- *β*1,*n*,j =*γ*1,*n*+*U*1,*n,j*

Where j represents subjects - j ∈[1,Npeople], Npeople= 30, and i represents time points - i ∈[0,1,3,6].

Time (here being the follow-up rounds) is treated as a discrete categorical variable. This allows us to model the effect of time on analyte progression without imposing parametric specifications on the shape of the relationship between time and analytes. Results for each biomarker are combined, and those with fixed effects with p-value ≤α=0.05 are considered significant, after false discovery rate (FDR) adjustment. While, traditionally, FDR adjustment is made via Bonferroni correction - α′=α/N - we opt here for the Benjamini & Hochberg (BH) FDR correction procedure (40) as it is more robust to false negatives and still adequately controls for family-wise error rate. This procedure is repeated for all time points, as well as for each time point in order to evaluate change between baseline and specific time points (D0 vs M1, D0 vs M3 and D0 vs M6). Results are quoted in log-fold changes, which is equal to the log-2 transform of the ratio between two elements (log_2_(A/B)).

### Clustering Analysis for QFT plasma samples

In order to further understand the structure of the biomarker data, we subjected the samples to an unsupervised clustering analysis as well as principal components analysis (PCA). The cytokines and biomarkers data were aggregated across all time points and all subjects, and analysed through K-means clustering. In both K-means and in PCA, we used the log-fold change in biomarker concentrations across all the follow-up timepoints to evaluate groupings and similarity between analytes. K-means clustering is an unsupervised learning algorithm that clusters data based on their similarity. It is carried out by evaluating the sum of squares distance between each analyte and the centroid of the K clusters, in terms of their log-fold change across the 6 months observation period of the study. At first, a number of clusters K is set. Each analyte is randomly assigned to one of the K clusters, then reassigned to the cluster that has the smallest distance from its centroid to that point. This process is reiterated until no further cluster reassignment occurs. K is chosen as the smallest integer which minimises the within-cluster sum of squares – a measure of within-cluster similarity.

## Data Availability

all data that is not in the supplements is available on request and RNAseq data was deposited to Gene Expression Omnibus (GEO) under accession number GSE158208.

## Study approval

The University of Cape Town Faculty of Health Sciences Human Research Ethics Committee approved these studies (HREC 245/2009 and 545/2010). All participants gave written informed consent prior to inclusion in the study, in accordance with the Declaration of Helsinki.

## Author contributions

KAW, RL, RJW conceived the experiments, RJW contributed resources, DL recruited participants and contributed samples. KAW, RL, NJ performed the experiments. DSL, CB, RL, GK, KAW performed data analysis. KAW and DSL wrote the draft manuscript and all authors contributed to manuscript revision, read and approved the submitted version.

## Acknowledgements

Research supported by Wellcome (104803, 087754, 203135), The South African National Research Foundation (443386), the European Union Horizon 2020 research and innovation programme under grant agreement No 643381 and The Francis Crick Institute which receives support from UKRI (FC001218), Cancer Research UK (FC001218) and Wellcome (FC001218). The Advanced Sequencing STP of the Francis Crick Institute performed the RNA sequencing experiments. We acknowledge Robert Goldstone and Philip East for helpful discussions related to the RNAseq experiments.

## Supplementary Figures

<u>Supplementary Figure 1.</u>

RNA sequencing analysis results, showing (A) Volcano plot visualising the differentially expressed genes identified in RNA sequencing analysis; and (B) Hallmark pathway analysis results.

<u>Supplementary Figure 2.</u>

Change in QFT plasma analytes compared to day 0, at 1M, 3M and 6M of ART. Minus log-transformed p-values for each analyte, resulting from the Limma analysis framework, after BH FDR correction at 1 Month, 3 Months and 6 Months of ART, compared to day 0. Sign represents direction of effect size (negative: decrease from D0 level, positive: increase from D0 level). Red lines: α significance thresholds.

<u>Supplementary Figure 3.</u>

Change in CD4 T cells expressing the chemokine receptors CXCR3, CCR4 and CCR6, determined by flow cytometry analysis in peripheral blood mononuclear cells in a subset of 25 patients from the same cohort, at day 0 and 6 months of ART.

## Supplementary Tables

**Supplementary Table 1.**
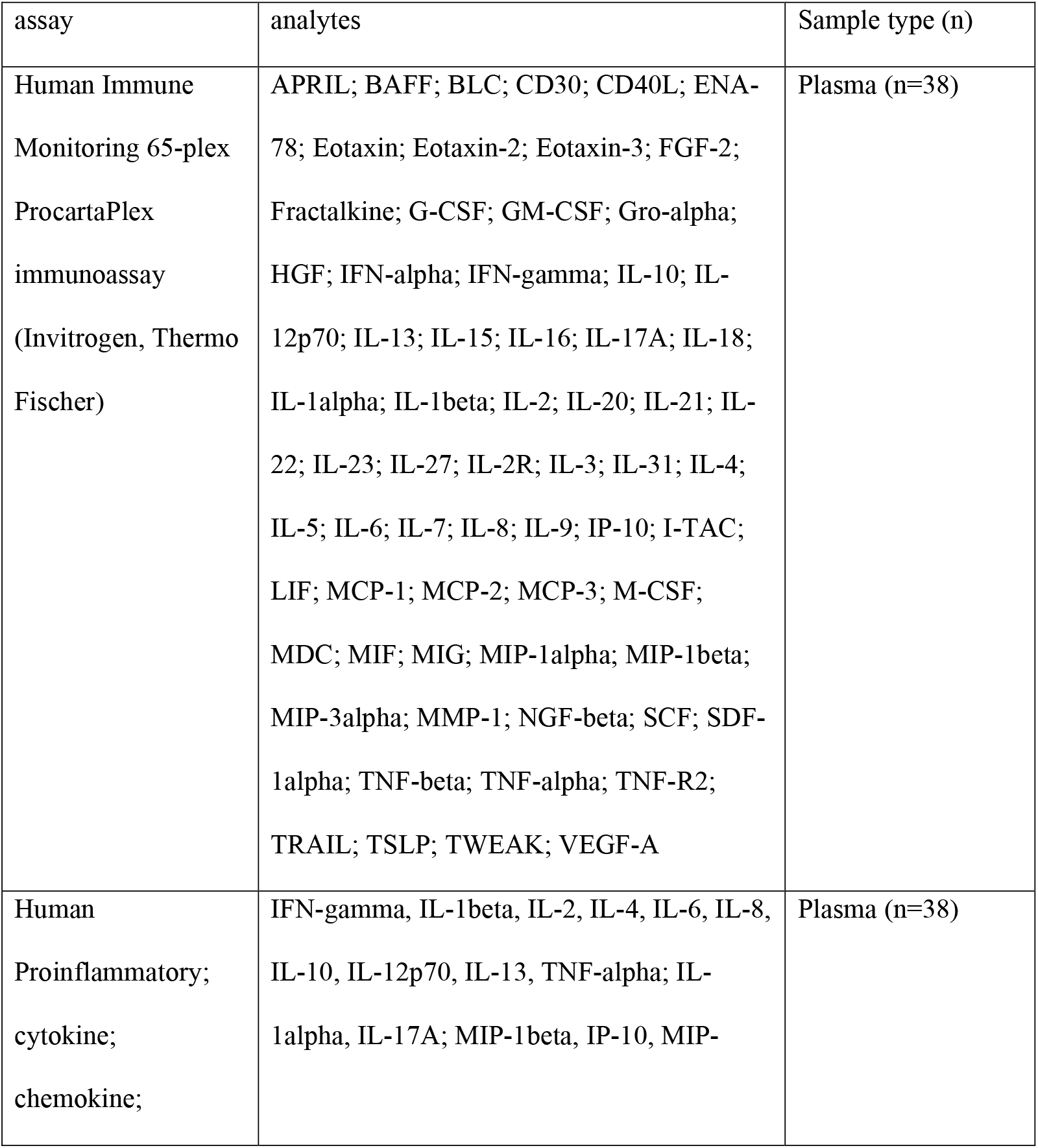
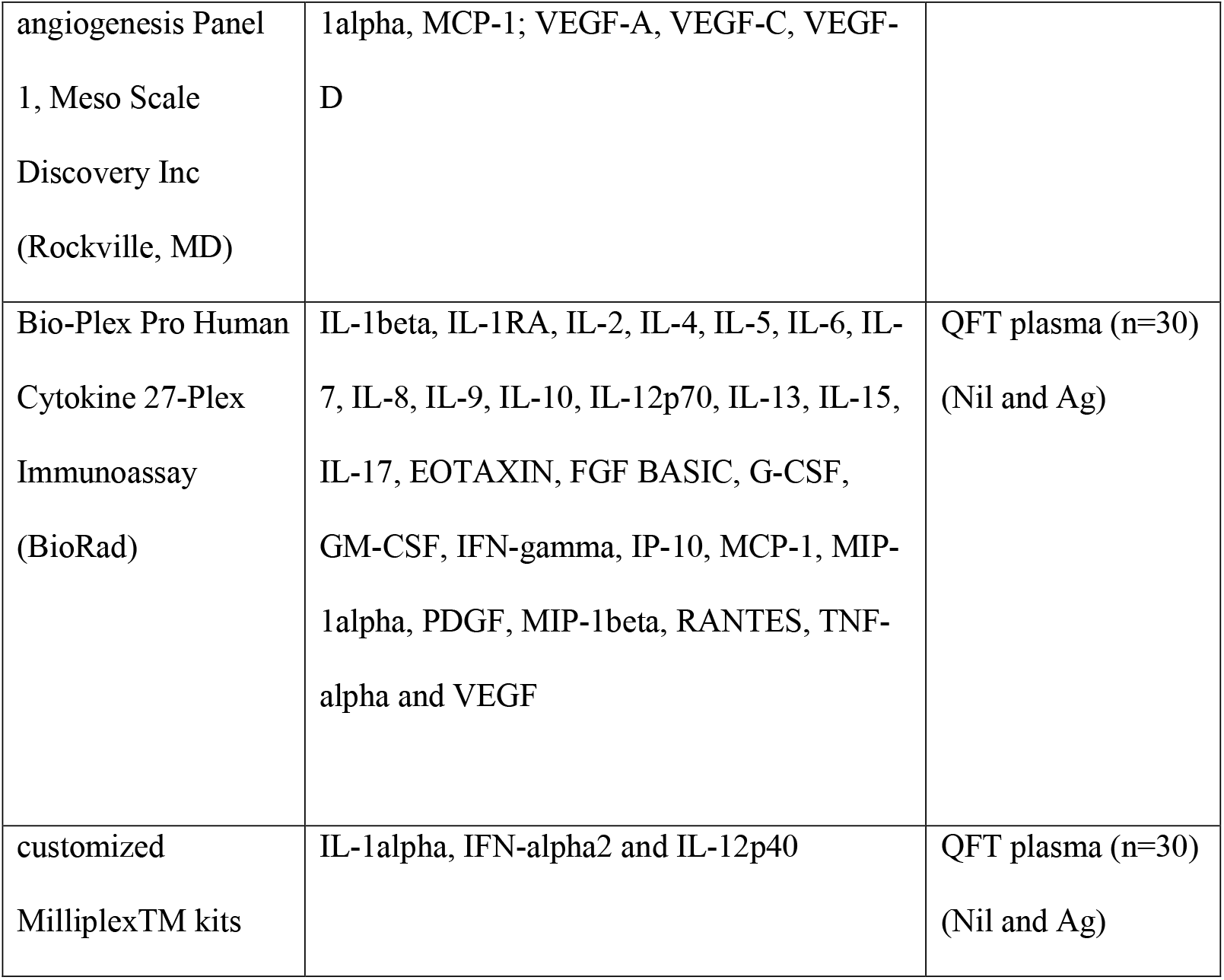
Analytes evaluated in the study.

**Supplementary Table 2.** Differentially expressed genes revealed by RNA sequencing This list is provided in an Excel spreadsheet.

**Supplementary Table 3.**
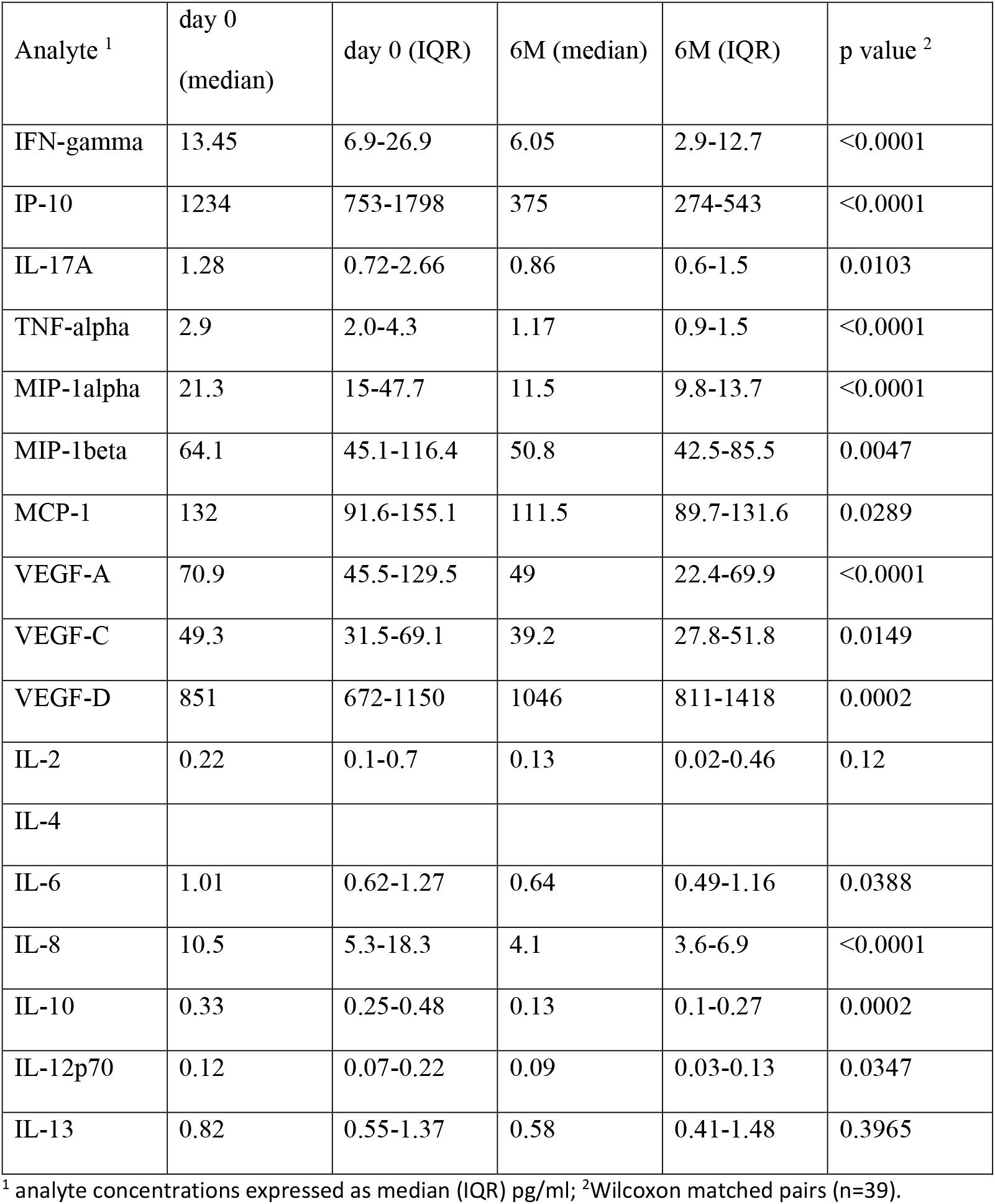
MSD analysis results.

**Supplementary Table 4.** HIV-1 viral load (VL) and CD4 T-cell counts (CD4) at each time point of the study in n=30 patients included in the QFT plasma analysis.

|  | D0 (Baseline) | Month 1 | Month 3 | Month 6 | P-value |
| --- | --- | --- | --- | --- | --- |
| VL<br>(mean (SD)) | 191,668.53<br>(315,605.35) | 506.47<br>(630.34) | 74.10 (66.02) | 507.41<br>(1,811.05) | <0.001 |
| CD4<br>(mean (SD)) | 203.83<br>(117.99) | 273.53<br>(149.09) | 285.67<br>(118.76) | 337.60<br>(137.63) | 0.002 |

**Supplementary Table 5.**
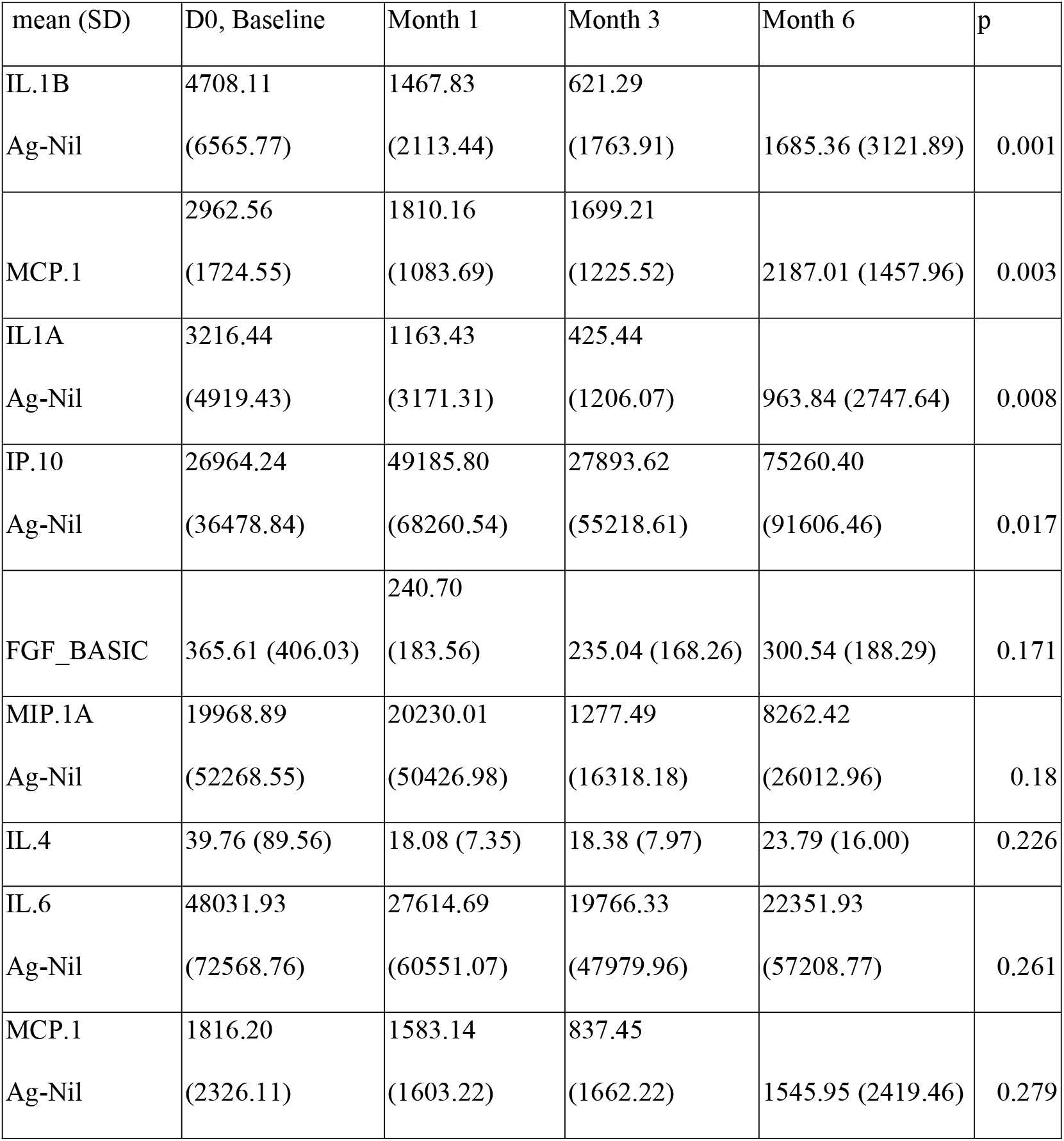
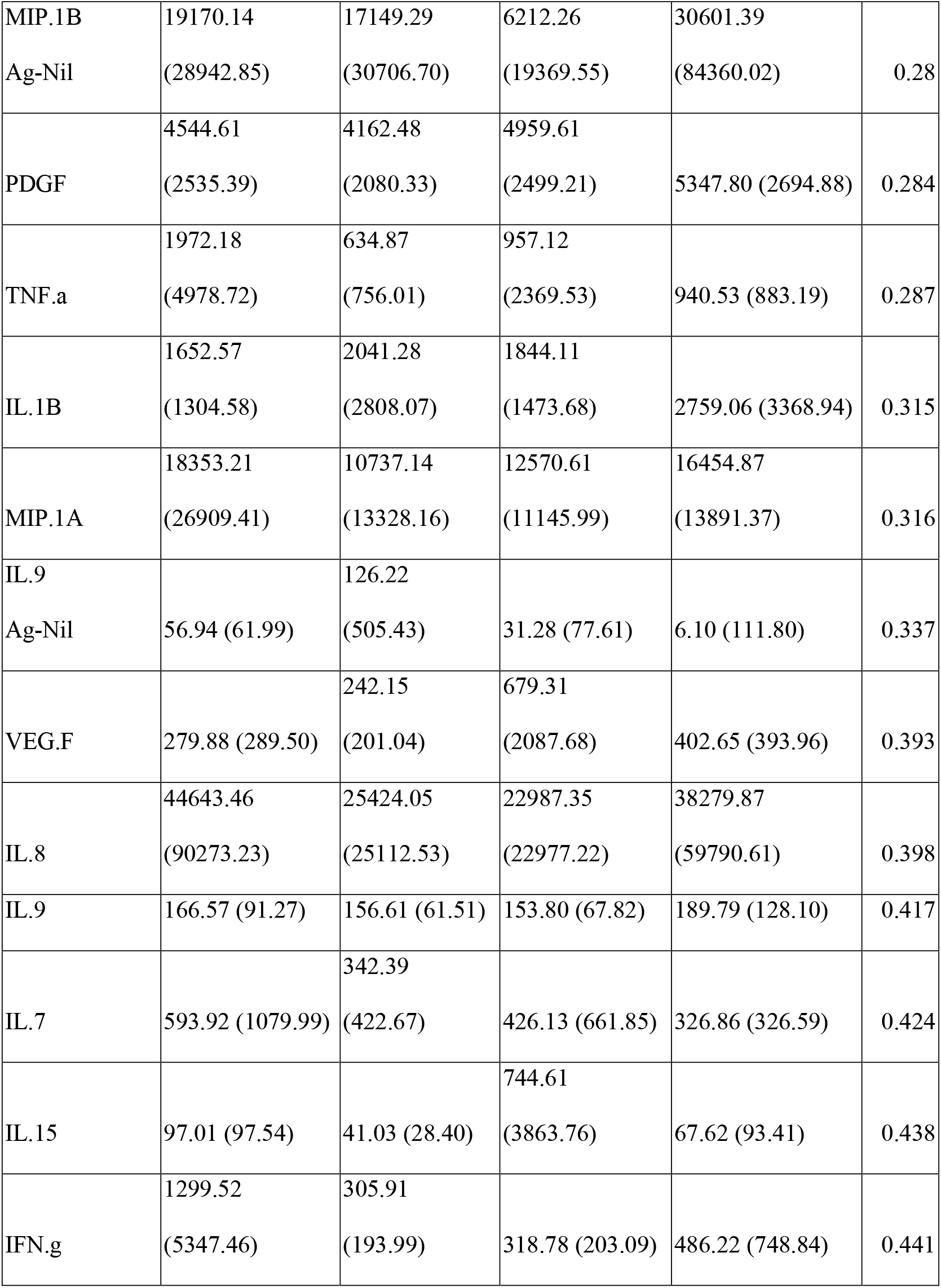
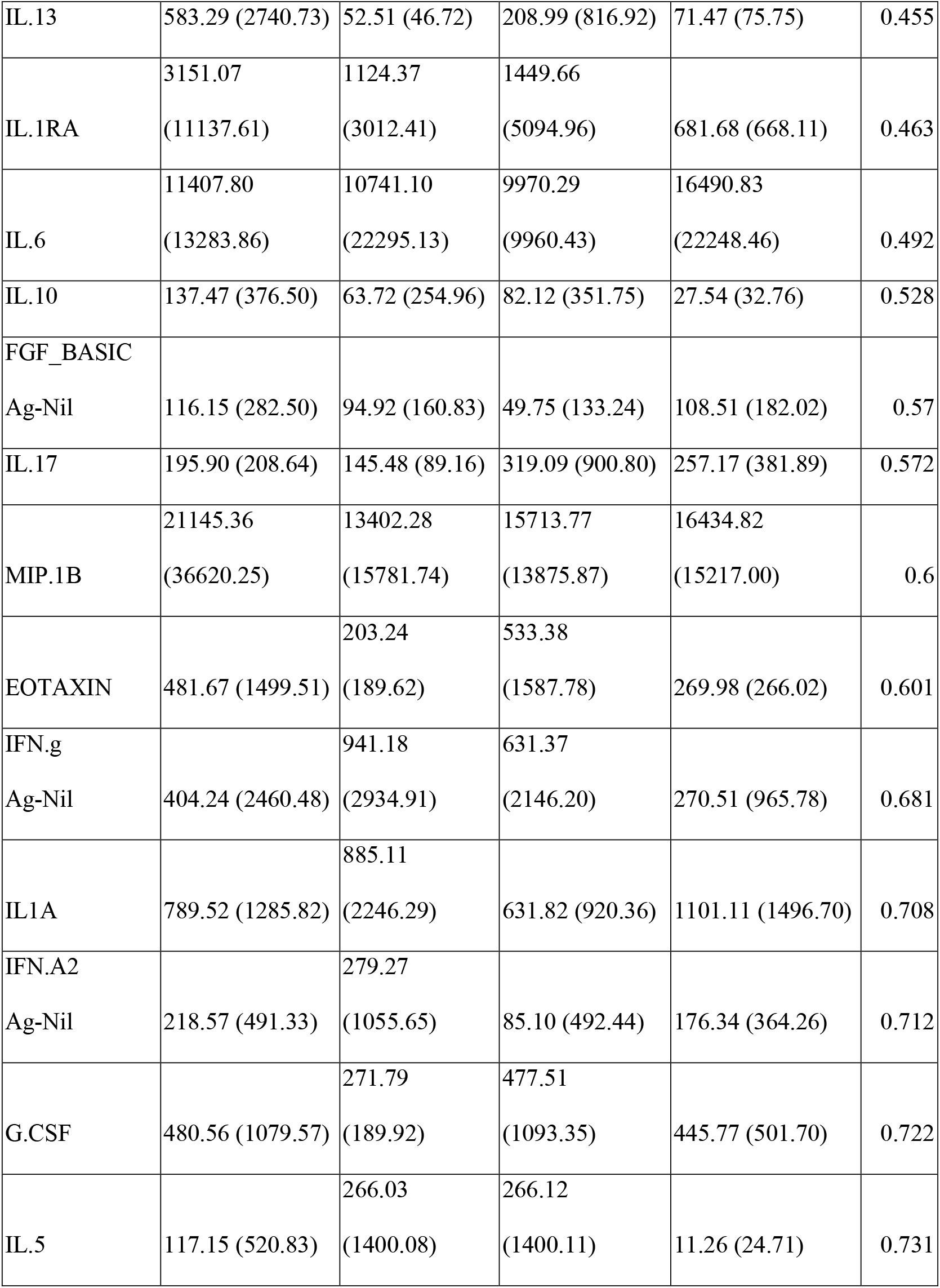
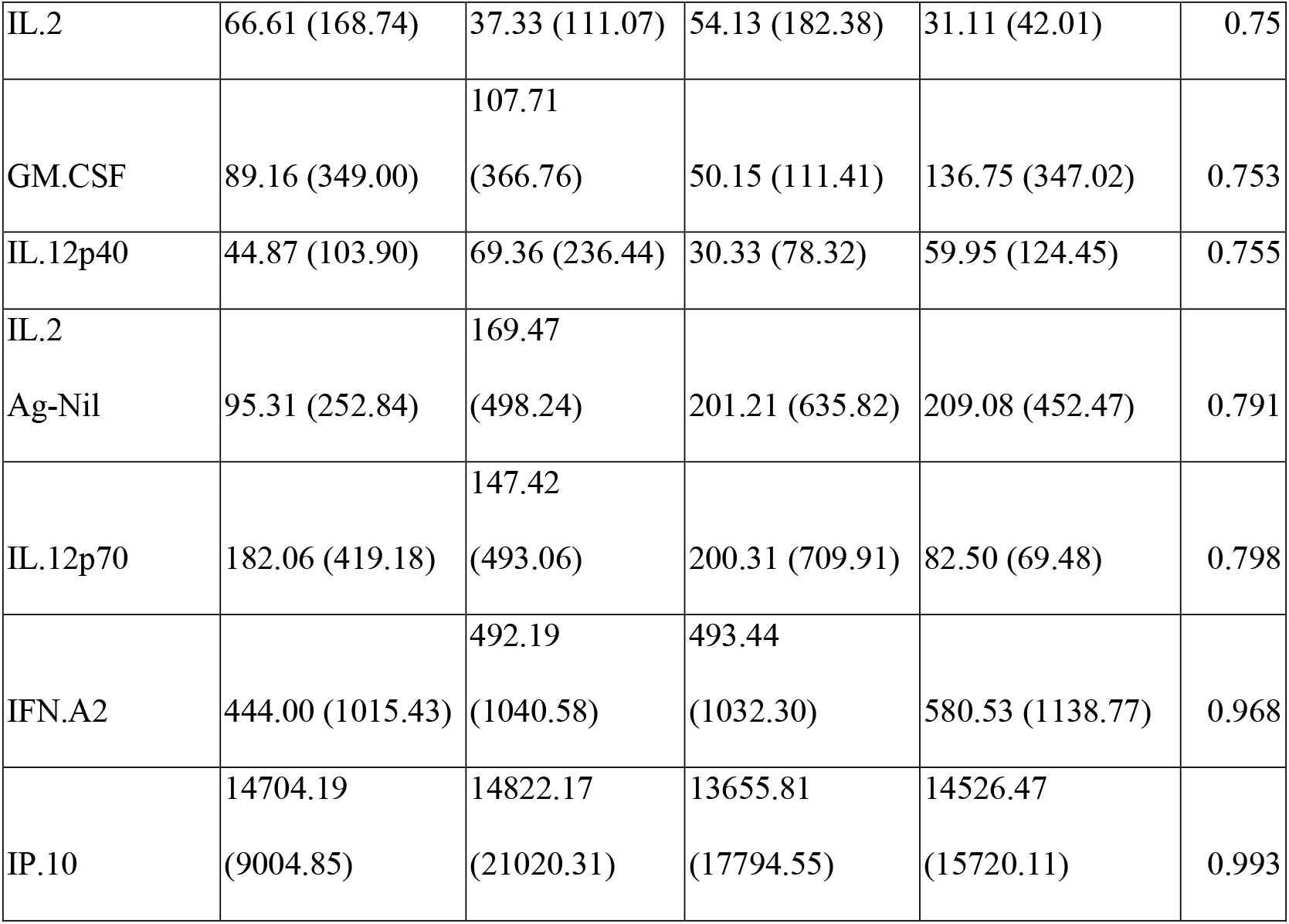
Mean (standard deviation, SD) in pg/ml for all analytes of interest in the study, across each time point in the n=30 patients included in the final analysis. P-value: p-value from group means test for each analyte, where H0 = no change in means across time point.

## Supplementary Figures

**Supplementary Figure 1A.**
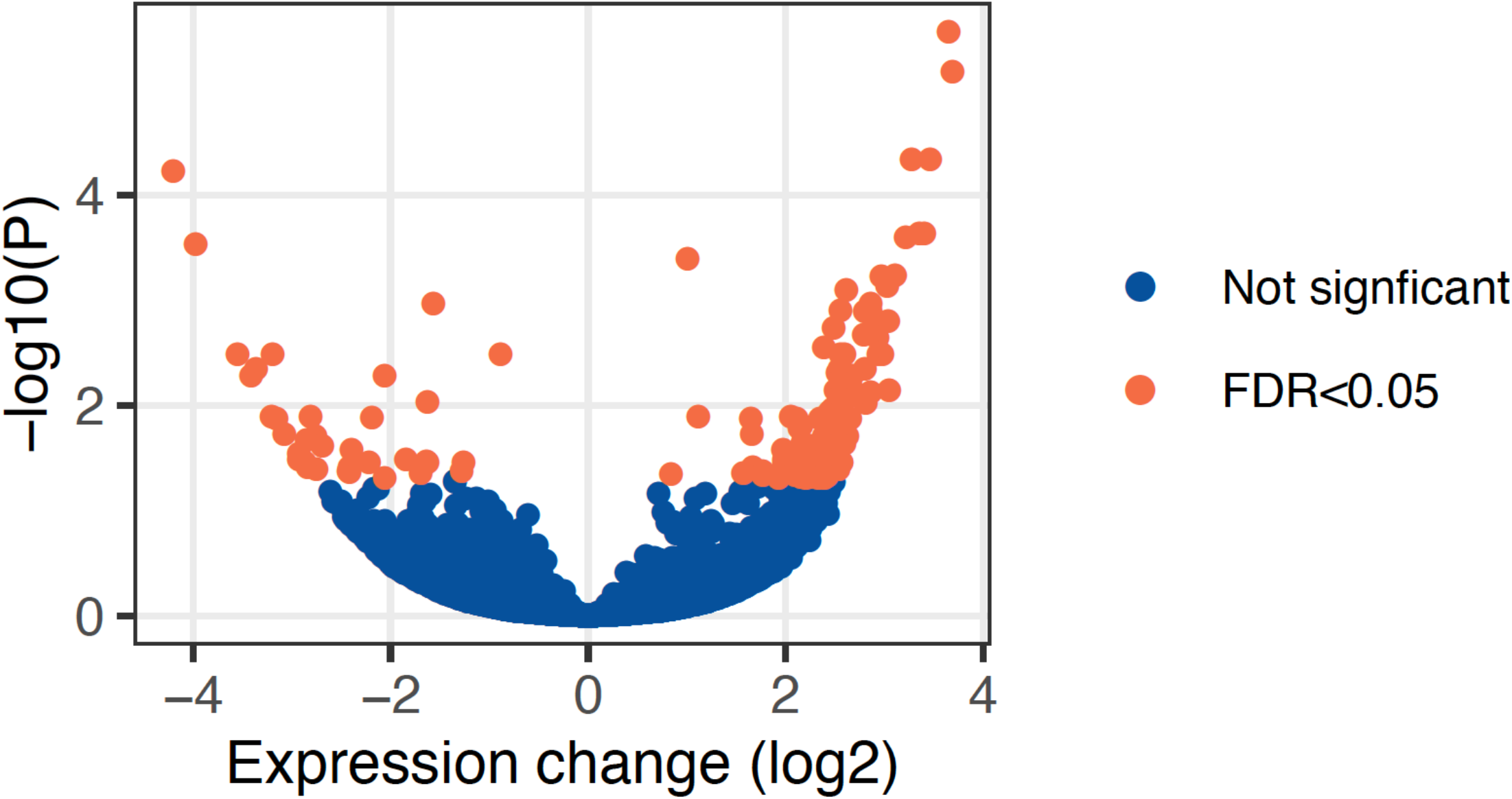
Volcano plot visualising the differentially expressed genes identified in RNA sequencing analysis.

**Supplementary Figure 1B.**
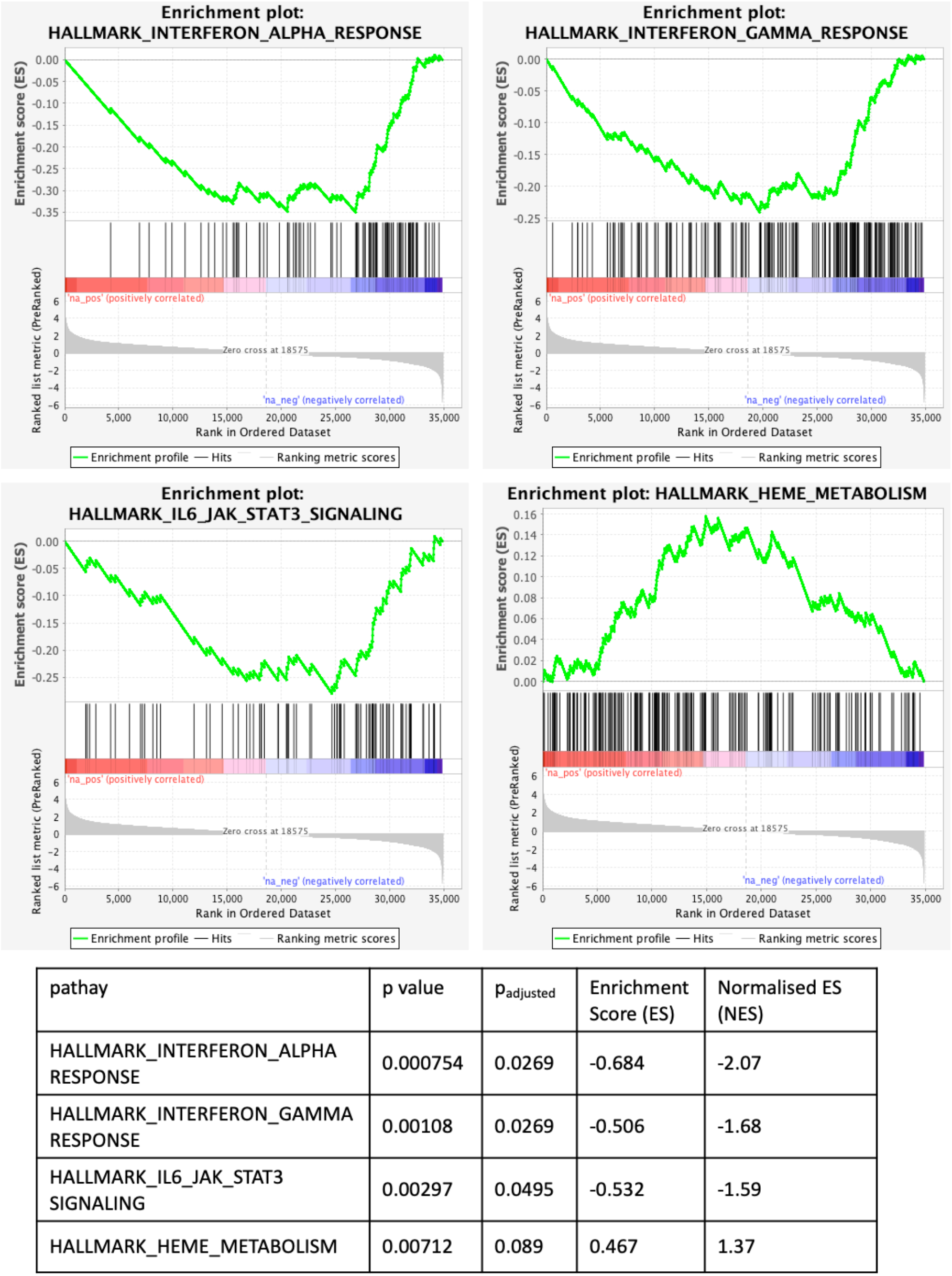
Hallmark pathway analysis results at 6M compared to D0.

**Supplementary Figure 2.**
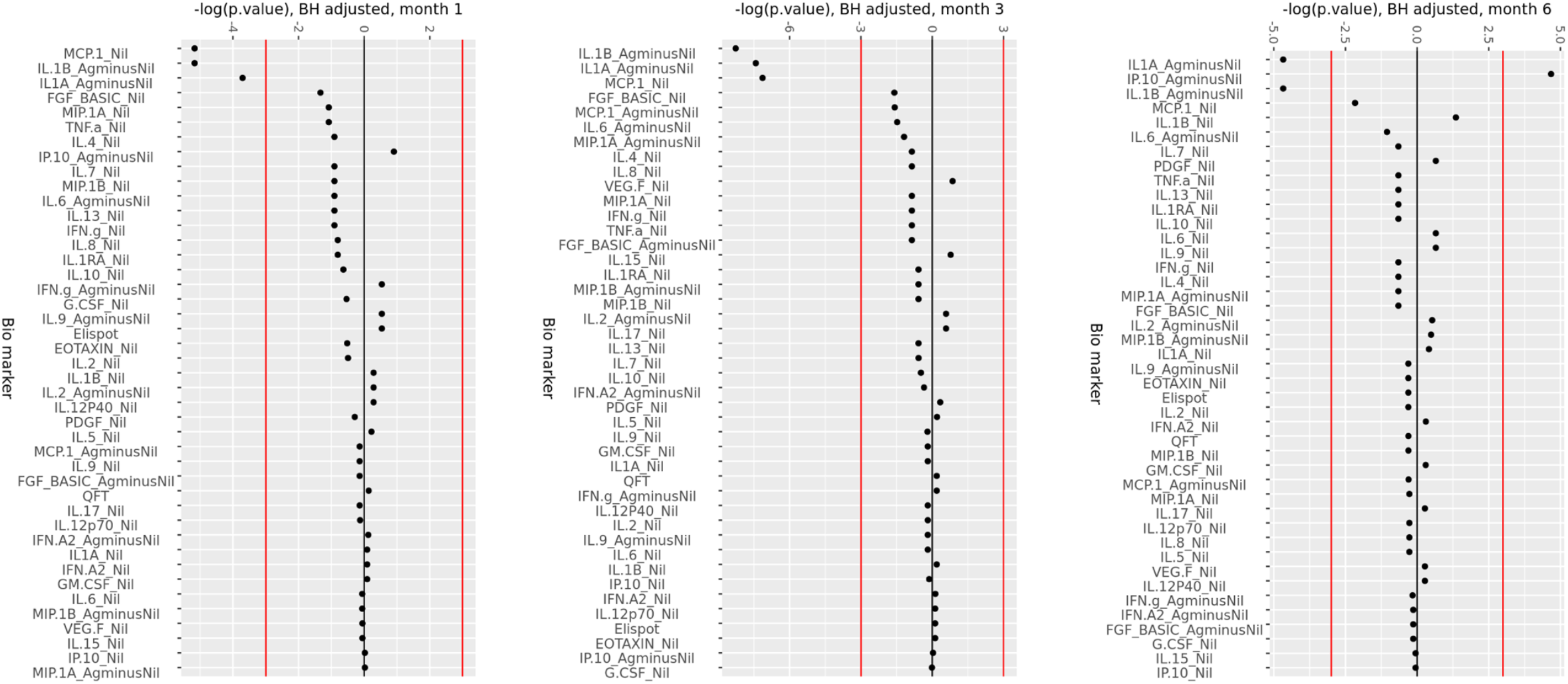
Change in analytes compared to day 0, at 1M, 3M and 6M of ART. Minus log-transformed p-values for each analyte, resulting from the Limma analysis framework, after BH FDR correction at 1 Month, 3 Months and 6 Months of ART, compared to day 0. Sign represents direction of effect size (negative: decrease from D0 level, positive: increase from D0 level). Red lines: α significance thresholds.

**Supplementary Figure 3.**
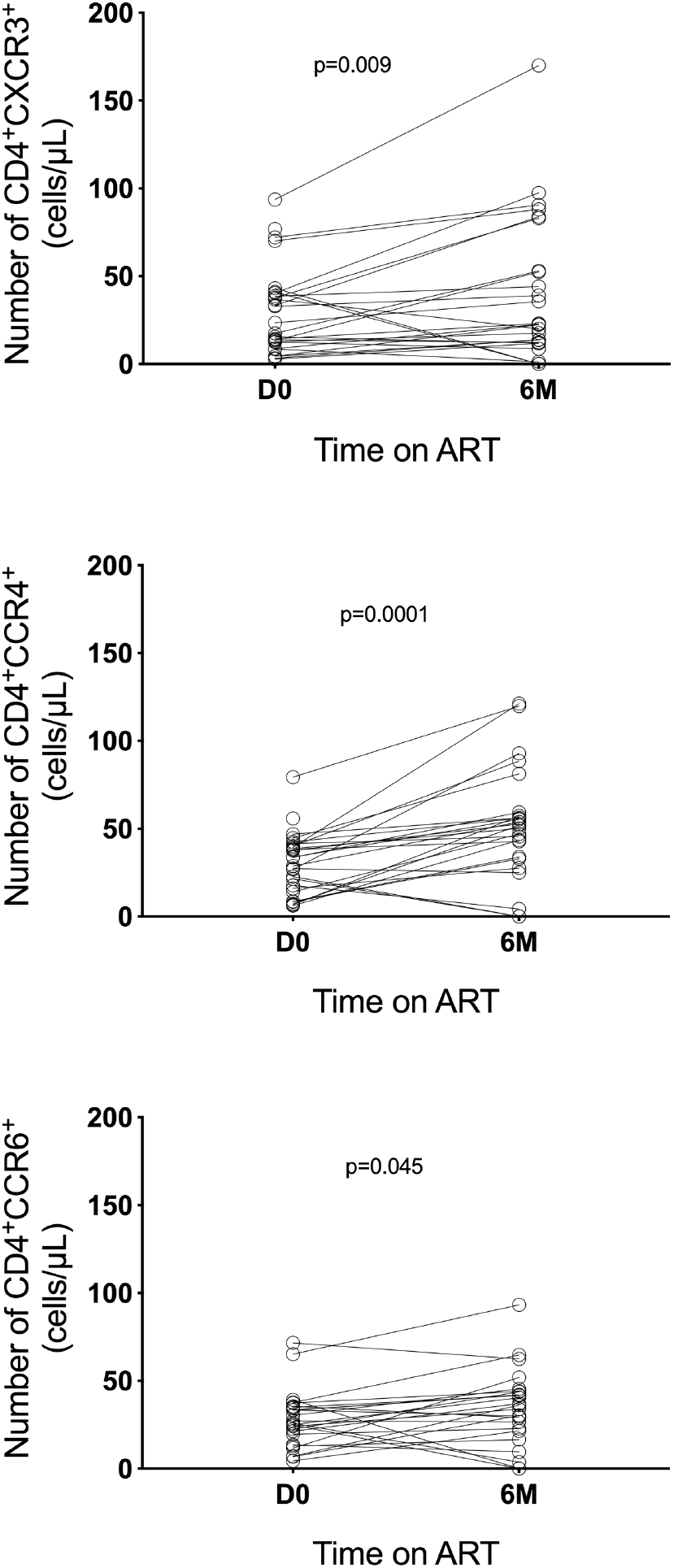
Change in CD4 T cells expressing the chemokine receptors CXCR3, CCR4 and CCR6, determined by flow cytometry analysis in peripheral blood mononuclear cells in a subset of 25 patients from the same cohort, at day 0 and 6 months of ART.

## Notes

### Competing Interest Statement

The authors have declared no competing interest.

